# A JOURNEY THROUGH RURAL COMMUNITY RADIOGRAPHY PRACTICE IN SOUTH-SOUTH NIGERIA: RADIOGRAPHERS’ PERCEPTION, WILLINGNESS AND BARRIERS

**DOI:** 10.1101/2022.09.07.22279697

**Authors:** Michael Promise Ogolodom, Awajimijan Nathaniel Mbaba, Beatrice Ukamaka Maduka, Chima Jude Iloka, Ikechukwu Felix Erondu, Uche Nathaniel Eja-Egwu, Nengi Alazigha, Egop Egop Brownson, Victor Kelechi Nwodo, Robert O. Akhigbe

## Abstract

**Introduction:** The lack of healthcare professionals and their retention in rural areas has become a serious concern to the health sector globally. Retaining health staff in rural areas has proven difficult as young professionals prefer urban postings. This study aimed to assess radiographers’ perception and willingness to work in rural areas of Rivers State.

**Method:** This cross-sectional questionnaire-based study was conducted among radiographers in Rivers State. The participants’ socio-demographic variables and their responses to willingness and perception to work in rural areas were obtained and analyzed.

**Results:** Only few 30% (n=12) of the respondents were willing to work in the rural areas of Rivers state. However, the majority of 95 %(n=38) of the respondents perceived extra payment as an incentive for them to be willing to work in the rural areas of this study location. Most 88 %(n=35) of the respondents perceived unfavorable working conditions in rural areas as a barrier to their willingness. A large proportion of 55 %(n=22) of the respondents stated that their marital status was a barrier to working in rural areas. More than half (85%, n=34) of the respondents perceived poor accommodation as a barrier to working in rural areas. The majority 88% (n=35) of the respondents stated that militant activities was a barrier to their willingness to work in the rural areas of Rivers State. There were statistically significant relationships between the evaluated respondent’s socio-demographic variables gender (χ^2^ = 48.000, p = 0.000), years of working experience (χ^2^ = 47.500, p = 0.000), marital status (χ^2^ = 84.966, p = 0.000) and age (χ^2^ = 76.758, p = 0.021) and their willingness to work in the rural areas of Rivers State.

**Conclusion:** The key findings suggest that the majority of the Radiographers were not willing to work in the rural areas of Rivers State. The reasons adduced for their strong disinclination were based on their perception of unfavorable working conditions in the rural areas. Nonetheless, they were of the opinion that financial inducement could influence their willingness to work in rural areas.

## INTRODUCTION

Radiographers are critical components of quality healthcare services and are in great demand worldwide. The radiographers provide both diagnostic and therapeutic services. In Nigeria, there is acute shortage and inappropriate distribution of radiographers. The availability and retention of healthcare professionals in rural areas is a challenge to the health sector globally [1]. Radiographers, like other health professionals prefer to work in urban areas due to higher incomes, good career opportunities, good infrastructures and social amenities [2,3].

Retaining health staff in rural areas has proven extremely difficult as young professionals prefer urban postings. Previous studies found that rural exposure, poor working conditions, low job satisfaction, political and ethnic problems, and sometime, civil strife and poor security in most rural areas, predispose young graduates to select urban centers [4,5]. A study conducted by Okeji et al[6] in Nigeria indicated that 27% of analyzed Radiographers including intern radiographers showed strong willingness to work in rural areas. A related study conducted by Khanagar et al[7] showed that 58% of dental interns were willing to work in rural areas of Riyadh, Saudi Arabia. Similarly, a study by Sharma et al[8] among medical students showed that 55% of the finalists were willing to work in rural communities.

Unemployment and opportunities influence regional labour migration because an unemployed worker is more likely to move with regional unemployment differentials encouraging mobility [9,10]. These factors which are not readily available within rural communities may then attract Radiographers to work within urban areas. People who move to urban areas are usually driven by their expectation of improved employment or earnings and desire to urbanize [11,12]. The rural/urban divide impacts negatively on access to both basic and comprehensive healthcare as well as skilled health professionals. In Nigeria, most Radiographers including radiography students prefer to work in urban areas whereas 54% of the population are rural dwellers [2,6].

Radiographers’ willingness to work in rural areas of Rivers State, Nigeria has not been previously studied. Hence the present study was carried out to assess the perception and willingness of radiographers towards working in rural areas of Rivers State, identify possible problems of working in rural areas and obtain possible solutions to assist in strategic health planning.

## METHODS

This was a cross-sectional survey design, which was conducted among radiographers in Rivers State. Rivers state is located in the South-South geopolitical zone of Nigeria with Port Harcourt as the administrative capital. The state is made up of rich and diverse cultures and ethnic groups such as Ogoni, Ikwerre, and Ijaw making up the greater population [13].

The study population comprised of qualified and licensed radiographers working in government and private-owned hospitals and diagnostic centers in Rivers state. Non-licensed radiographers and x-ray technicians were excluded from the study. Ethical approval (NAU/FHST/2021/RAD34) for this study was obtained from the Research Ethical Committee of the Faculty of Health Sciences and Technology, College of Health Sciences, Nnamdi Azikiwe University, Nnewi Campus, Anambah State, Nigeria. The aim of the study was adequately to the participants and their consent was duly sought and obtain. Their privacy and confidentiality were guaranteed and they were at liberty to withdraw from the study at any time without any harm.

The sample size for the study was determined using the statistical formula for unknown population used by previous study Charan and Biswas[14].

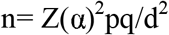

Where;

n=expected sample size

Zα = significant level usually set at 95% confidence level, Zα is 1.96 (two sided)

p= portion of the population with similar attributes under study = 60% (0.6)

d= margin of error tolerated or absolute error = 15.2% (0.152)

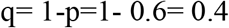

Therefore; n= 1.96^2^ ×0.6×0.4 / 0.152^2^

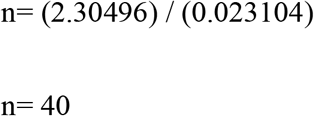

A convenience sampling technique was used to select the 40 participants. The consent of the participants were duly sought and obtained using written informed consent form.

A 23 items self-completion questionnaire was the instrument for data collection. The questionnaire contained twenty three (23) questions divided into three (3) sections. Section I: elicited information on radiographer’s socio-demographic variables, section II was concerned with general questions on radiographer’s willingness to work in rural communities and section III consisted of questions evaluating the barriers militating against radiographer’s willingness to work in rural areas.

### Validity and reliability Test

A pilot study was carried out with 10 questionnaires distributed among radiographers before the commencement of this study and the Cronbach alpha reliability test was computed. The questionnaire had an acceptable internal consistency (Cronbach’s alpha = 0.87).

The validity of the questionnaire was calculated using the Index of term Objective Congruence (IOC) method used by previous authors [15-17]. This was done by computing the index of item-objective congruence (IOC). Based on the index parameters, an IOC score greater than 0.6 was assumed to show adequate content validity, and all the scores obtained in this study for all the items of the questionnaire after IOC analysis was greater than 0.6

The questionnaire was in electronic and hardcopy versions. The electronic version was issued using Whatsapp group of Association of Radiographers of Nigeria (ARN)-Rivers State chapter and participant’s feedback was retrieved electronically. The hardcopy version was distributed by the researchers using one-on-one method and retrieved immediately after completion. This study was conducted from December 2021 to February 2022.

The data collected were analyzed using the Statistical Package for Social Sciences software (SPSS) version 21.0(IBM Corp, Armonk, NY, 2012). The result of the data analyzed was represented using frequency tables and percentages. Inferential statistics was done using Chi-square to evaluate the association between participant’s socio-demographic variables and their willingness to work in rural areas of Rivers State. The level of statistical significance was set at p-value less than 0.05.

## RESULT

Out of 40 respondents, 70% (n=28) were males while females accounted for 30% (n=12). A good number 67.5% (n=27) were within the age bracket of 21-30 years and the least 5%(n=2) were < 20 years. Single respondents were highest in number 67.5 %(n=27). The majority 80% (n=32) had only B.Sc degree and only 2.5 %(n=1) was a Ph.D holder. More than half 65% (n=26) had 0-5 years working experience and the least 2.5 %(=1) had 11-15 years working experience(Table 1).

**Table 1:** Frequency and percentage distributions of the socio-demographic variables.

| Variables | Classification | Frequency N=40 | Percent |
| --- | --- | --- | --- |
| Gender | Female | 12 | 30% |
|  | Male | 28 | 70% |
| Class Total |  | 40 | 100% |
| Marital status | Married | 13 | 32.5% |
|  | Single | 27 | 67.5% |
| Class Total |  | 40 | 100% |
| Qualification | B.Sc | 32 | 80% |
|  | M.Sc | 7 | 17.5% |
|  | Ph.D | 1 | 2.5% |
| Class Total |  | 40 | 100% |
| Years of Experience | 0-5 | 26 | 65% |
|  | 6-10 | 13 | 32.5% |
|  | 11-15 | 1 | 2.5% |
| Class Total |  | 40 | 100% |
| Age | <20 | 2 | 5% |
|  | 21-30 | 27 | 67.5% |
|  | 31-40 | 8 | 20% |
|  | 41-50 | 3 | 7.5% |
| Class Total |  | 40 | 100% |

Only few 30% (n=12) of the respondents were willing to work in the rural areas of Rivers state. However, the majority 95 %(n=38) of the respondents perceived extra payment as an incentive for them to be willing to work in the rural areas of this study location. Out of 40 respondents, only 25% (n=10) had lived in the rural areas. A greater number 53% (n=21) of the respondents had no previous experience working in rural areas (Figure 1).

**Figure 1:**
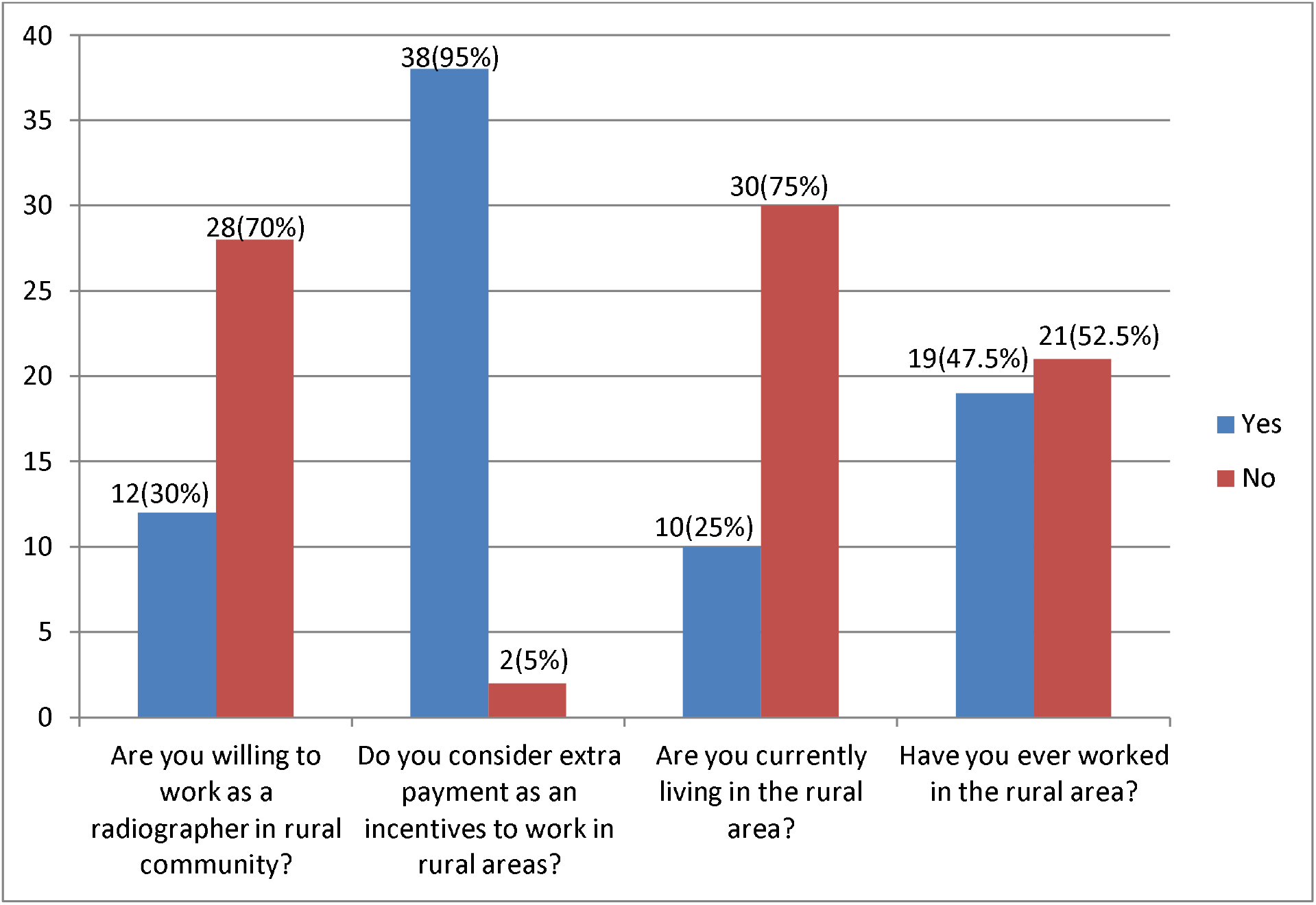
Frequency and percentage distributions of the radiographer’s responses to questions on their willingness to work in rural areas.

The responses to questions on barriers to radiographer’s willingness to work in rural areas of Rivers State as noted in table 3 are; most of the respondents 87.5%(n=35) perceived unfavorable working conditions in the rural areas as a barrier to their willingness. A large proportion 55 %(n=22) of the respondents stated that their marital status was a barrier to working in rural areas. More than half (85%, n=34) of the respondents perceived poor accommodation as a barrier towards working in rural areas. The majority 87.5% (n=35) of the respondents agreed that militant activities was a barrier to their willingness to work in the rural areas of Rivers State (Table 2).

**Table 2:**
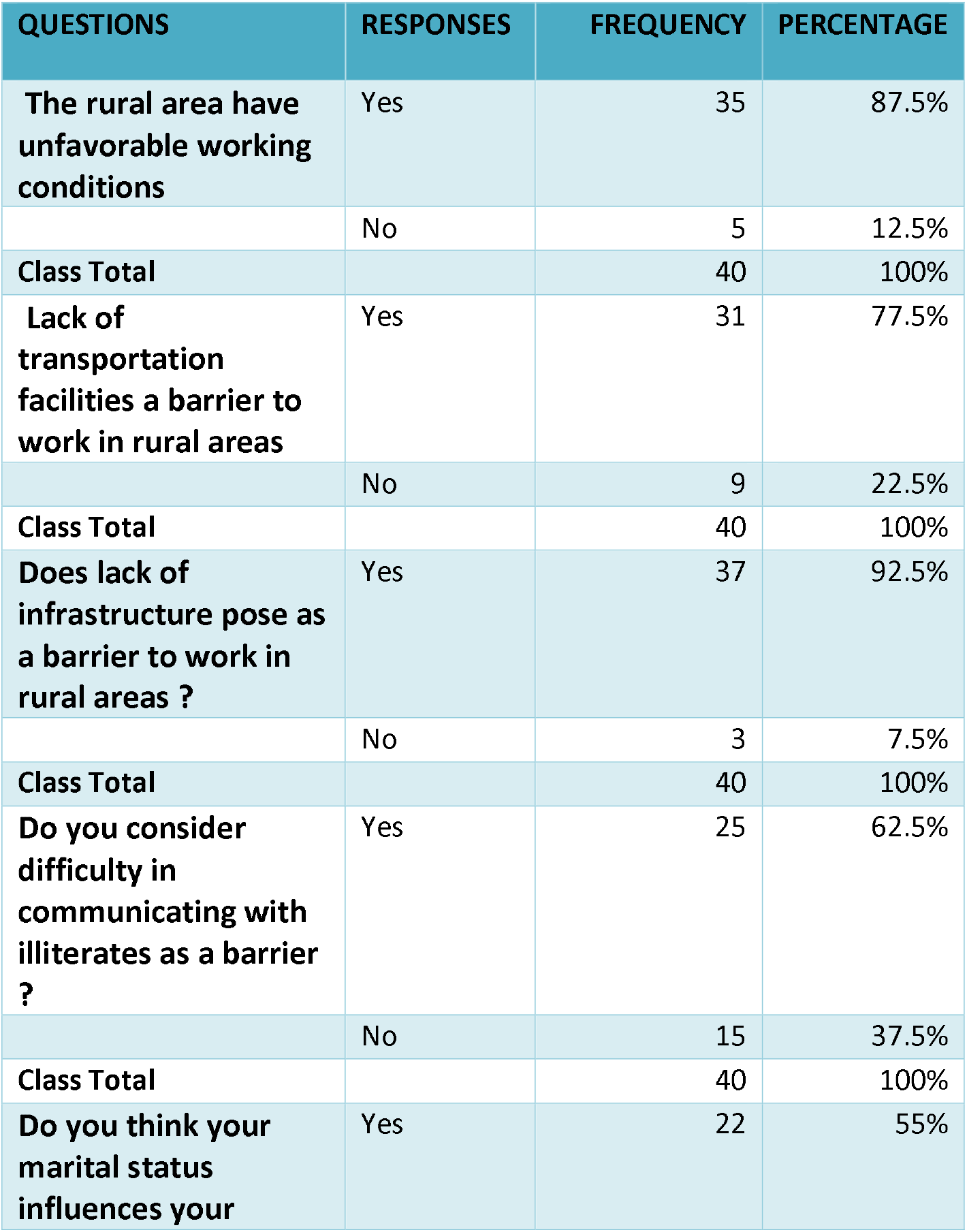
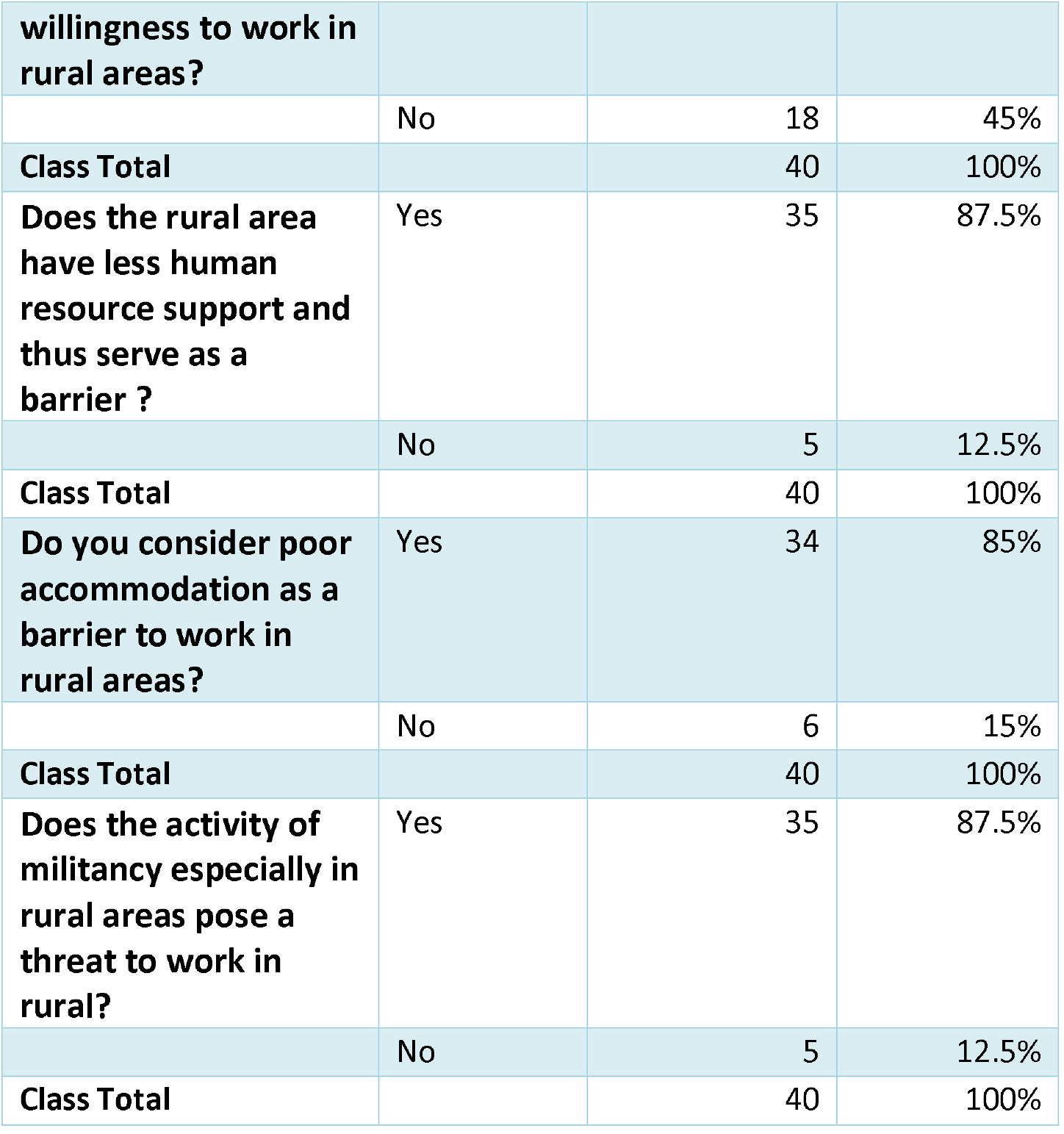
Frequency and percentage distributions of the radiographer’s responses questions on barriers to their willingness to work in rural areas.

**Table 3:**
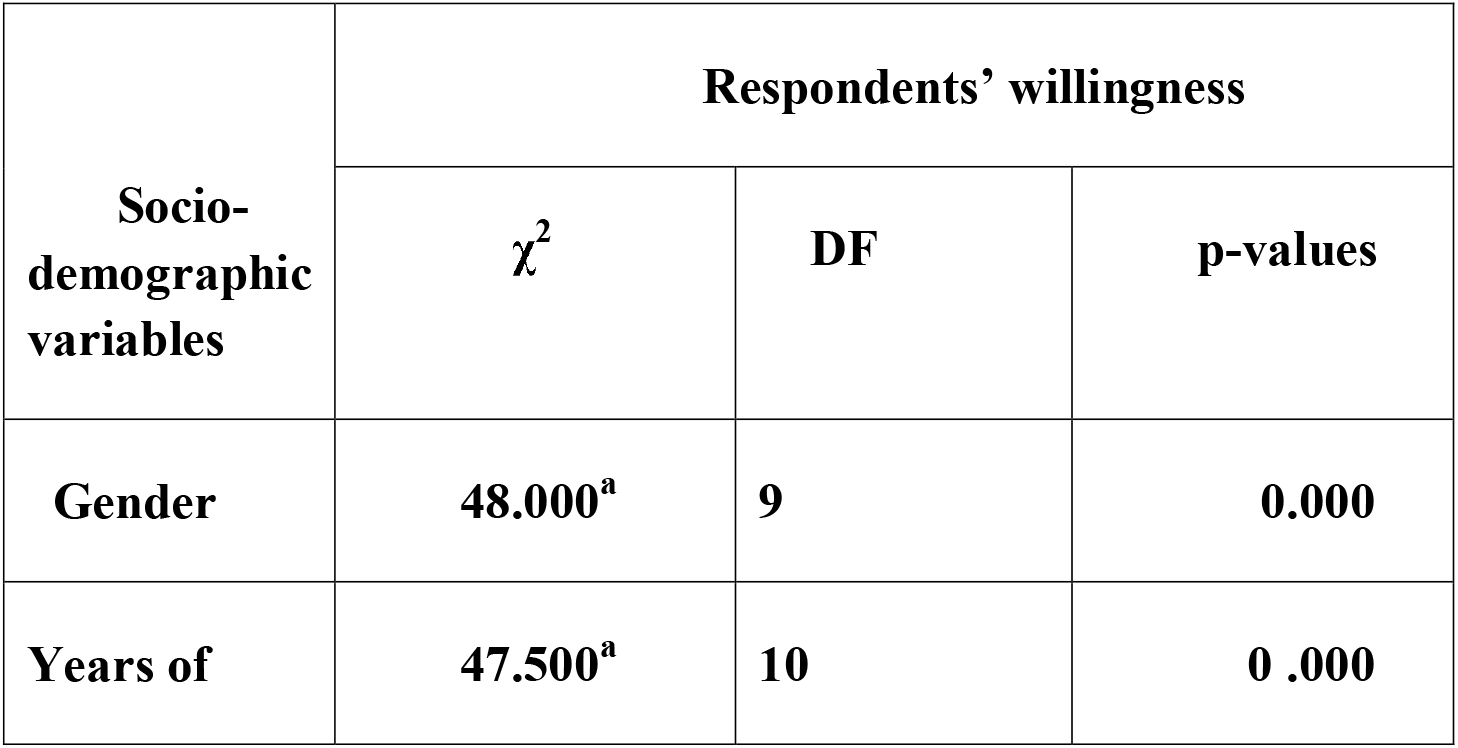
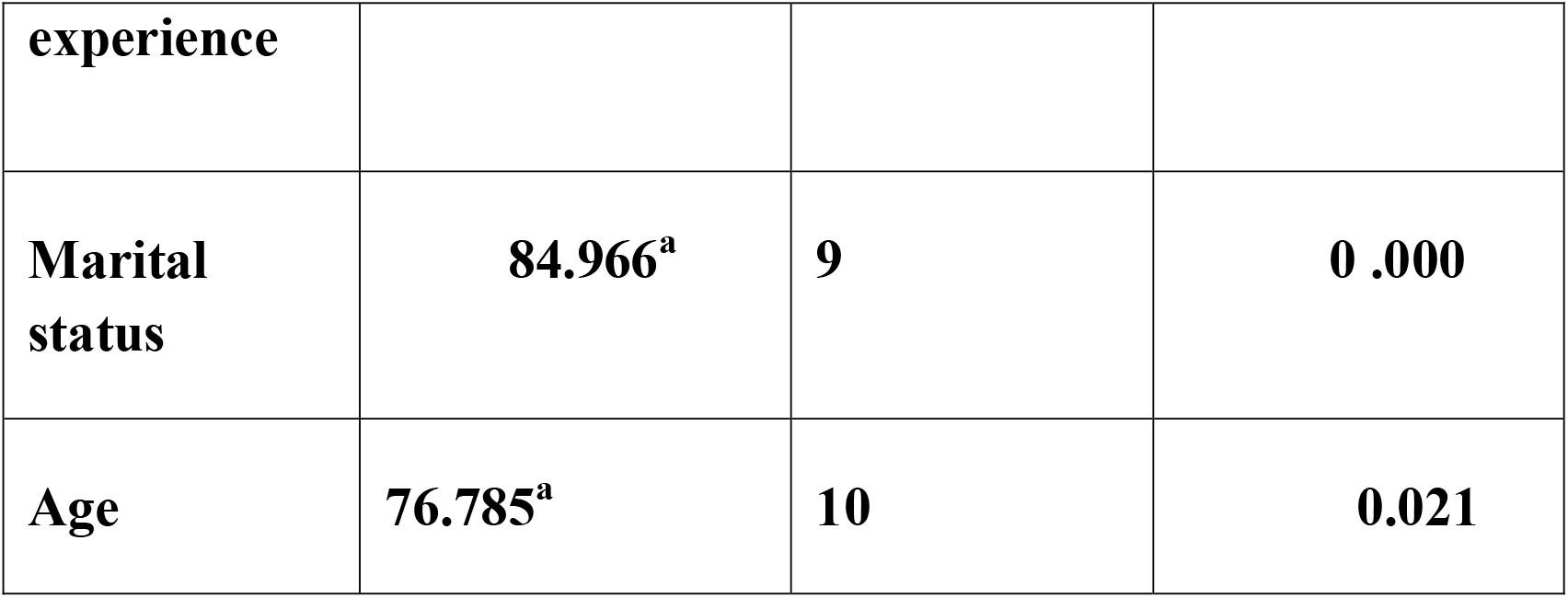
Associations between radiographer’s gender, years of experience and marital status and their willingness to work in rural areas.

There were statistically significant relationships between the evaluated respondent’s socio-demographic variables gender (χ^2^ = 48.000, p = 0.000), years of working experience (χ^2^ = 47.500, p = 0.000), marital status (χ^2^ = 84.966, p = 0.000) and age (χ^2^ = 76.758, p = 0.021) and their willingness to work in the rural areas of Rivers State (Table 3).

## DISCUSSION

Radiographers are critical in the provision of quality healthcare services to the population and they render diagnostic and therapeutic services, and are invaluable for the monitoring of treatment outcomes. Increased utilization of facility-based health services becomes more visible in some rural communities due to the launching of imaging services [18,19]. Imaging has also been proved to impact management decisions [20,21].

We found that very few of the respondents were positive and willing to work in the rural areas of Rivers state. This is inconsistent with the findings of similar studies conducted by Thammatacharee et al[22] in Thai among medical, dental and pharmacy graduates, Khanagar et al[7] in Riyadh, Saudi Arabia among dental interns, Okeji et al[6] among final year radiography students in Southeast, Nigeria and Anzenberger et al[23] among health workers in Ukraine, which reported that the majority of the participants were positive and willing to work in the rural communities areas of their study locations. The discrepancies in our findings could be ascribed to the different sample sizes studied, geographical locations of the studies as well as the compositions of the study population. According to Anzenberger et al[23], the participants were interested in working in rural areas as long as opportunities align with their individual expectations.

Despite the fact that there was poor willingness to work in rural areas of Rivers State among the radiographers in our study, the majority of the respondents perceived extra payment as an incentive for them to be willing to work in the rural areas as the topmost factor that would motivate to work in rural areas. This may be due to the fact that most radiographers would like to make more earning to enable them invest in the rural areas, and also cater for their family. This finding is in harmony with those of a previous study carried out by Okeji et al[6] in Southeast, Nigeria among final year radiography students, which also documented remuneration as the topmost motivating factor to work in rural areas among their participants. According to Okeji et al[6], they noted that their participants felt that as upcoming professionals, they were more interested in high earning to assist them start life and alleviate their family challenges. Contrary to the finding of this index study, Khanagar et al[7] conducted a study in Riyadh, Saudi Arabia among dental interns and reported close proximity to hometown as the topmost factor that motivated their participants’ willingness to work in rural areas. This difference in our findings could be ascribed to the different sample size used in our different studies.

We identified multi-factors, which form barriers to radiographers’ willingness to work in rural areas of Rivers state and these include but not limited to lack of infrastructures, unfavorable working conditions in the rural areas, marital status, poor accommodation and militancy activities in rural areas. Lack of infrastructure was the topmost factor, followed by unfavorable working condition and militant activities in these rural areas. This finding implies that the respondents were dissatisfied with the organizational structure of hospitals/diagnostic centers, and security situation in rural areas of the state. This may be so because health priorities in rural areas are more focused on prevention of infectious diseases [24]. Consequently imaging may be less prioritized. This may explain the paucity of imaging equipment in rural areas. In addition, the imaging equipment is less sophisticated, older, and in poor functional state [21]. These challenges/barriers to our participant’s willingness to work in rural area of the state may result to fewer/ radiographers working in the rural areas of the state.

There were statistically significant relationships between the evaluated respondent’s socio-demographic variables gender, years of working experience, marital status and age, and their willingness to work in the rural areas of Rivers State. These findings imply that sex, years of working experience, marital status as well as the age of the respondents has so much significant influence on their willingness to work in rural areas of the state. Married couples who were already residing in the urban centres may find it difficult to relocate to the rural areas. This may be so because they have to consider their family conditions as well as the challenges of lack social amenities that are commonly associated with rural areas. Also, young radiographers may find it difficult to work in the rural areas because a good number of them may be willing to advance their education and also wish to work with sophisticated equipment. The imaging equipment is less sophisticated, older, and in poor functional state in most of the rural areas [21].

## CONCLUSION

The key findings suggest that the majority of the Radiographers were not willing to work in the rural areas of Rivers State. The reasons adduced for their strong disinclination were based on their perception of unfavourable working conditions in the rural areas. However, radiographers’ remuneration is a major deciding factor that would enhances the willingness to work in rural communities, while Social ties and working conditions are barriers to urban to rural migration among radiographers in River state

To attract young energetic Radiographers to the rural areas, specific policy intervention should be established, including provision of basic amenities and improving the security situations in the rural areas. The government is therefore urged to provide state of the art imaging facilities in rural areas and improve on the pay package to attract more radiographers in the rural areas

## Data Availability

All data produced in the present study are available upon reasonable request to the authors

## Ethical Approval

**The approval (**NAU/FHST/2021/RAD34) was

## Acknowledgement

**Not applicable**

## Author Contributions

All authors have read and approved the manuscript. Each author participated sufficiently in this submission and the roles of the authors are: MPO, CJI, ANM and BUM were the main researchers, drafted the manuscript, responsible for data capturing, presentation of results, MPO, BUM, IFE, UNE, NA, EEB, VKN and ROA carried out the interpretation of results and also gave recommendations on the review of literatures, and provide critical comments on the research work.

## References

1. Sidibe′ CS, Toure′ O, Broerse JEW, Dieleman M. Rural pipeline and willingness to work in rural areas: Mixed method study on students in midwifery and obstetric nursing in Mali. PLoS ONE 2019; 14(9): e0222266.

2. Ossai E N, Anyanwagu U C, Azuogu B N, Uwakwe K A, Ekeke N, Ibiok N C. Perception about Working in Rural Area after Graduation and Associated Factors: A Study among Final Year Medical Students in Medical Schools of Southeast Nigeria. Journal of advances in Medicine and Medical Research 2015; 8(2): 192–205. DOI: 10.9734/BJMMR/2015/16937

3. Dussault G, Franceschini MC. Not enough there, too many here: understanding geographical imbalances in the distribution of the health workforce. Hum Resour Health. 2006; 4:12. DOI: 10.1186/1478-4491-4-12.

4. Agyei-Baffour Kotha SR, Johnson JC, Gyakobo M, Asabir K, Kwansah J et al. Willingness to work in rural areas and the role of intrinsic versus extrinsic professional motivations - a survey of medical students in Ghana. BMC Medical Education 2011; 11:56

5. Kruk ME, Johnson JC, Gyakobo M, Agyei-Baffour P, Asabir K, Kotha SR, Kwansah J, Nakua E, Snowg RC, Dzodzomenyo M. Rural practice preferences among medical students in Ghana: a discrete choice experiment. Bull WHO 2010; 88:333–341.

6. Okeji MC, Ugwuanyi DC, Adejoh T. Radiographers’ willingness to work in rural and underserved areas in Nigeria: a survey of final year radiography students. Journal of Association of Radiographers of Nigerian 2014; 28 (1): 6–10

7. Khanagar SB, Alfaran KM, Alenazi YB, Aloqayli AM, Alsahhaf AH, Alotaibi FR et al. Willingness and Perception of Dental interns towards working in rural areas in Riyadh, Saudi Arabia: A cross-sectional study. Journal of Clinical and Diagnostic Research 2020; 14(8):Z001–Z005.

8. Sharma V, Gupta N, Rao NC. Perception towards serving rural population amongst interns from dental colleges of Haryana. Journal of Clinical and Diagnostic Research 2014;8(9):ZC31–32. doi: 10.7860/JCDR/2014/8978.4832.

9. Rabe B, Taylor M. Differences in Opportunities? Wage, Employment and House-Price Effects on Migration. RePEc. 2010: Available from: https://www.researchgate.net/publication/46469066_8_Differences_in_Opportunities_Wage_Unemployment_and_House-Price_Effects_on_Migration.

10. Pissarides CA, Wadsworth J. Unemployment and the inter-regional mobility of labour. Economic Journal 1989; 99(397):739–755. https://doi.org/10.2307/2233768.

11. Glomm G. A model of growth and migration. Canadian Journal of Economics 1992; 25(4):901–922. https://doi.org/10.2307/135771.

12. Lyu H, Dong Z, Roobavannan M, Kandasamy J, Pande S. Rural unemployment pushes migrants to urban areas in Jiangsu Province, China. Palgrave Commun. 2019; 5(92): https://doi.org/10.1057/s41599-019-0302-1

13. About Rivers State Government. 2021. Available from https://www.riversstate.gov.ng/about/. Retrieved on 21st December 2021

14. Charan J, Biswas T. How to calculate sample size for different study designs in medical research?. Indian Journal of Psychological Medicine 2013;35:121–126.

15. Ogolodom MP, Mbaba AN, Alazigha N, Erondu OF, Egbe NO, et al. (2020) Knowledge, Attitudes and Fears of HealthCare Workers towards the Corona Virus Disease (COVID-19) Pandemic in South-South, Nigeria. Health Sciences Journal 1:002

16. Mbaba AN, Ogolodom MP, Abam, R, Akram M, Alazigha N et al. Willingness of Health Care Workers to Respond to Covid-19 Pandemic in Port Harcourt, Nigeria. Health Sciences Journal 2021; 15 (1), 802.

17. Turner R C, Carlson L. Indexes of item-objective congruence for multidimensional items. International Journal of Test 2003;3:163–171. https://doi.org/10.1207/S15327574IJT0302_5.

18. Bashour H, Hafez R, Abdulsalam A (2005). Syrian women’s perceptions and experiences of ultrasound screening in pregnancy: Implications for antenatal policy. Reprod Health Matters 13(25): 147–154. doi: 10.1016/s0968-8080(05)25164-9

19. Kimberly HB, Murray A, Mennicke M, Lieplo A, Lew J, Bohan JS et al. Focused martenal ultrasound by midwives in rural Zambia. Ultrasound Medical Biology 2010; 36(8):1267–72. DOI: 10.1016/j.ultrasmedbio.2010.05.017.

20. Stein W, Katunda I, Butoto C. A two-level ultrasonographic service in a maternity care unit of a rural district hospital in Tanzania. Tropical Doctor 2008; 38(2):125–126. PMID: 18453515 DOI: 10.1258/td.2007.070045

21. Kawooya MG, Pariyo G, Malwadde EK, Byanyima R, Kisembo H. Assessing the diagnostic Imaging needs for five selected hospitals in Uganda. Journal of Clinical Imaging Sciences 2011; 1:53. DOI: 10.4103/2156-7514.90035

22. Thammatacharee N, Suphanchaimat R, Wisaijohn T, Limwattananon S, Putthasri W. Attitudes toward working in rural areas of Thai medical, dental and pharmacy new graduates in 2012: A cross-sectional survey. Human Resources and Health. 2013; 11:53. https://doi.org/10.1186/1478-4491-11-53.

23. Anzenberger P, Popov SB, Ostermann H. Factors that motivate young pharmacists to work in rural communities in the Ukraine. Rural and Remote Health 2011; 11: 1509. https://doi.org/10.22605/RRH1509

24. Cooke JG. Public Health in Africa: A report of the CSIS Global Health Policy Centre. Centre for Strategic and International Studies. Journal Public Health in Africa 2009; 1(1) DOI: 10.4081/jphia.2010.e8

